# Association of serum platelet-to-lymphocyte ratio levels with the risk of stent thrombosis and long-term prognosis in patients undergoing percutaneous coronary intervention with and without type 2 diabetes mellitus: a large-scale prospective cohort study

**DOI:** 10.1101/2023.03.14.23287277

**Authors:** Yanjun Song, Zhangyu Lin, Jining He, Kongyong Cui, Chenxi Song, Rui Zhang, Boqun Shi, Qiuting Dong, Kefei Dou

**Author notes:** **Correspondence to:** <u>Kefei Dou, MD, PhD,</u> Cardiometabolic Medicine Center, Department of Cardiology, Fuwai Hospital, National Center for Cardiovascular Diseases, State Key Laboratory of Cardiovascular Disease, Chinese Academy of Medical Sciences and Peking Union Medical College, No. 167, Beilishi Road, Xicheng District, Beijing 100037, China, And <u>Qiuting Dong, MD, PhD</u> Cardiometabolic Medicine Center, Department of Cardiology, Fuwai Hospital, National Center for Cardiovascular Diseases, State Key Laboratory of Cardiovascular Disease, Chinese Academy of Medical Sciences and Peking Union Medical College, No. 167, Beilishi Road, Xicheng District, Beijing 100037, China. Yanjun Song, Zhangyu Lin, and Jining He contributed equally to this article. Kefei Dou and Qiuting Dong contributed equally as senior authors.

## Abstract

**Background:** The platelet-to-lymphocyte ratio (PLR) is a promising inflammatory biomarker contributing to the development of atherosclerosis and type 2 diabetes mellitus (T2DM). Therefore, this study aims to document the value of PLR in predicting adverse events in patients undergoing percutaneous coronary intervention (PCI) with and without T2DM.

**Methods:** This study consecutively enrolled 8831 patients who received PCI, and we divided them into 4 groups according to the PLR level and glycemic metabolic statuses (PLR-low/high with non-T2DM, PLR-low/high with T2DM). The endpoints were major adverse cardiovascular and cerebrovascular events (MACCE) and stent thrombosis. Multivariate COX regression analysis was used to determine the association.

**Results:** During the 2.4-year follow-up, a total of 663 (7.5%) MACCE and 75 (0.85%) stent thrombosis were recorded. Results showed that patients with high PLR levels presented a significantly higher risk of MACCE (HR: 1.26, 95%CI: 1.07 to 1.47, *P* = 0.005) and stent thrombosis (HR: 2.29, 95%CI, 1.39 to 3.79, *P* = 0.001) when compared with the PLR-low group. When focused on patients with T2DM, the PLR-high group showed a significantly higher risk of MACCE (HR: 1.53, 95%CI: 1.17 to 2.00, *P* = 0.002) and stent thrombosis (HR: 3.79, 95%CI, 1.62 to 8.86, *P* = 0.002). However, these associations were not significant in patients without T2DM.

**Conclusions:** For the first time, PLR is documented as a great predictor for the poor prognosis and high incidence of stent thrombosis in patients with CAD, especially in those with T2DM.

**CLINICAL PERSPECTIVE:** *What Is New?:*

- In patients undergoing percutaneous coronary intervention (PCI), high serum platelet-to-lymphocyte ratio (PLR) is associated with poor prognosis and a high risk of stent thrombosis.
- When focusing on patients with different glycemia statuses, we found that the predictive value of PLR for poor prognosis and high risk of stent thrombosis was significant in patients with type 2 diabetes mellitus (T2DM), but not in patients without T2DM.
- Among patients with different PLR levels and glycemia statuses, those combined with both high PLR levels and T2DM showed the highest risk of poor prognosis and stent thrombosis.
- In stent thrombosis events, very late stent thrombosis is the most common type in patients with high PLR.

*What Are the Clinical Implications?:*

- Patients undergoing PCI with high PLR levels should be monitored closely.
- Residual inflammation should be focused among patients with coronary heart disease after revascularization to improve the long-term prognosis.

## 1. Introduction

High inflammatory state has been regarded as a key promotor in the pathogenesis of coronary artery disease (CAD) ^1^. Rigorous studies have reported that diverse inflammatory actors modulated various pathologic roles, such as driving atheroprogression and inciting plaque destabilization, thereby contributing to the following clinical performances like angina pectoris or acute coronary syndrome ^2^. As for patients undergoing percutaneous coronary intervention (PCI), the elevated inflammatory burden was suggested to be associated with aggravated endothelial dysfunction and microvascular obstruction, which finally contributed to PCI-related myocardial injury and high long-term all-cause mortality ^3, 4^. Therefore, precise indicators evaluating inflammatory burden and their predictive value for prognosis in patients with CAD hold great attention.

Platelet-to-lymphocyte ratio (PLR) was first described as a prognostic marker in periampullary cancer in 2008 ^5^ and demonstrated to reflect inflammation and thrombosis in various cancers in the following studies. Recently, PLR has shown its value in evaluating inflammatory burden and indicating poor prognosis in several cardiovascular diseases like ST-elevation myocardial infarction (STEMI) ^6, 7^, heart failure ^8^, stable angina pectoris, chronic total occlusion ^9^ and in patients with an implantable cardioverter defibrillator ^10^. Platelet-induced proinflammatory cytokines release and atherothrombotic progression might be the potential mechanisms illustrating its value of reflecting inflammatory state in cardiovascular diseases ^11^. Besides, the prognostic value of PLR in glycemic metabolism disease has also been reported, as high PLR levels in patients with type 2 diabetes mellitus (T2DM) showed a high incidence of complications like diabetic nephropathy and cognitive decline ^12, 13^.

Therefore, the relationship between PLR and clinical outcomes in CAD patients with different glycemic metabolism statuses after PCI, whose related research is still limited, raised interest. In this study, we prospectively enrolled 8831 patients undergoing PCI, and divided them into different groups according to their glycemic statuses and PLR levels, aiming to document the predictive roles of PLR for the prognosis in these patients.

## 2. Methods

### 2.1. Study design and population

In this prospective cohort study, 10724 patients who underwent PCI from January 2013 to December 2013 were consecutively screened. PCIs were all performed at Fuwai Hospital, Chinese Academy of Medical sciences. Adult patients (>18 years old) who were treated with drug-eluting stents (DES) implantation were eligible for this research. Exclusion criteria include a combination of hematological disorders, active tumors, infection, immune-suppressant or steroid drug application, and missing baseline data. After that, 8831 patients were ultimately enrolled in this study **(Figure S1)**. Platelet and lymphocyte counts were collected from one blood sample before the PCI procedural, and PLR was calculated using the formula of plasma platelet count (10^9^/L) to plasma lymphocyte count (10^9^/L) ratio ^11^. To investigate the prognostic value of PLR in CAD under different glycemic states, enrolled patients were firstly divided into two groups: PLR-Low (n=4419) and PLR-High (n=4412), and further allocated into four groups: PLR-H-T2DM (n=1263), PLR-L-T2DM (n=1404), PLR-H-Non-T2DM (n=3149), PLR-L-Non-T2DM (n=3015). The optimal PLR cut-off was defined as the median of all participants (PLR=107, interquartile range: 84.7-135.5), and T2DM was defined as a history of T2DM from the on-admission medical record, receiving oral hypoglycemic agents or insulin, fasting glucose level ≥126 mg/dL, or hemoglobin A1c (HbA1c) level ≥ 6.5%. All patients received a clinical follow-up with a median time of 2.4 years after the operation. The clinical status was evaluated at 1, 6, and 12 months in the first year after discharge, and annually assessed in the following years by telephone interview.

The research process was following the Declaration of Helsinki and was approved by the Institutional Review Board of Fuwai Hospital, Chinese Academy of Medical sciences. All subjects provided informed written consent for long-term follow-up before intervention.

### 2.2. Data collection and definitions

Data on variables of interest were prospectively collected in all participants, which comprised demographic data (age, sex, BMI, comorbidities, past histories, smoking or not), on-admission diagnosis, physical/imaging/laboratory examination, target lesion/vessel information, PCI procedure and medicine therapy after PCI/at discharge. Comorbidities collected in this study included diabetes mellitus, hypertension (more than twice on different days by systolic blood pressure ≥ 140 mmHg and/or diastolic blood pressure ≥ 90 mmHg during the baseline hospitalization or known hypertension with antihypertensive medication), and dyslipidemia (plasma total cholesterol [TC] ≥ 5.2 mmol/L, triglyceride [TG] ≥ 1.7 mmol/L, low-density lipoprotein cholesterol [LDL-C] ≥ 3.4 mmol/L, high-density lipoprotein cholesterol [HDL-C] < 1.0 mmol/L, and/or known dyslipidemia receiving lipid-lowering therapy). Past histories included previous myocardial infarction (MI), revascularization (PCI/coronary artery bypass grafting [CABG]), peripheral artery disease (PAD), and stroke. Chronic coronary syndrome (CCS) or acute coronary syndrome (ACS) was diagnosed according to on-admission clinical presentation. As for coronary disease, left main artery disease (LM, ≥50% stenosis in LM coronary artery), three-vessel disease ( ≥ 50% in all three main epicardial coronary arteries), and lesion characteristics (chronic total occlusion [100% occlusion of a native coronary artery for more than 3 months with thrombolysis in myocardial infarction (TIMI) flow grade of 0], bifurcation, calcification, and syntax score [estimated using an online calculator (http://www.syntaxscore.com/)]) were recorded by two independent interventionists at Fuwai hospital. The number of the treated vessel and implanted DES during PCI and procedural success (the residual stenosis ≤ 30% and grade 3 TIMI flow without apparent dissection) or not were collected as procedural data. Medicine therapies after PCI include dual antiplatelet therapy (DAPT), GPIIb/IIIa receptor antagonists, β-blocker, statins, and nitrate.

### 2.3. Laboratory examination

Blood samples were collected from patients before PCI and stored in −80 °C refrigerators until the test. The process of laboratory examination was performed at the core laboratory in Fuwai hospital, according to the standard operation guideline. In this study, routine blood tests (Hgb, white blood cells [WBC], lymphocyte, platelet), lipid profiles (TG, TC, HDL-C, and LDL-C), creatinine, high-sensitivity C-reactive protein (hs-CRP) and glycosylated hemoglobin A1c (HbA1c) were presented.

### 2.4. Procedures

PCIs were all performed by experienced interventionalists in Fuwai hospital. Before PCI, patients all received a loading dose of DAPT (300 mg of aspirin and 300 mg of clopidogrel the day or ticagrelor 180 mg) and heparin treatment with an initial bolus of 70-100 IU/kg (before the procedure) with an additional dose of 1000 IU administered per hour (during the procedure). The access of intervention was radial or femoral artery. Angiography was performed in all participants, and the application of adjunctive examinations (intravascular ultrasound and optical coherence tomography) and devices (balloon and stent) choice depended on the operators.

### 2.5. Outcomes

The primary endpoints of this study were major adverse cardiovascular and cerebrovascular events (MACCE, a composite of all-cause death, MI, stroke, and target vessel revascularization [TVR]). The secondary outcomes were the individual components of MACCE and stent thrombosis. MI was diagnosed according to the third universal definition of MI ^14^ and TVR referred to any repeat revascularizations of the treated vessel in PCI. Stroke was defined as cerebrovascular events (ischemic or hemorrhagic) confirmed by two neurologists and stent thrombosis was defined as a thrombotic occlusion of a coronary stent (defined according to Academic Research Consortium criteria) ^15^. The time definitions of acute stent thrombosis (AST), subacute stent thrombosis (SAST), late stent thrombosis (LST), and very late stent thrombosis (VLST) were 0-24 hours, 24 hours to 30 days, 30 days to 1 year and longer than 1 year after stent implantation, respectively ^16^. The events above were all confirmed by at least two independent specialists of cardiology, and the disagreement was resolved by consulting a third experienced cardiologist.

### 2.6. Statistical analysis

Categorical variables were presented as “number (%)”. Continuous variables were described as “mean ± standard deviation (SD)” when they were normally distributed. Two-sample T-test was used to assess whether there was a statistical difference in continuous variables when they were normally distributed, otherwise, the Mann-Whitney U test was used. χ² test was applicated to test the differences in categorical variables, although Fisher’s exact test was used for the small sample size. Kaplan-Meier (K-M) plots and Cox proportional hazards regression models were used for survival analysis, which was based on the time from operation to primary outcomes. KM plots for PLR-L vs. PLR-H, PLR-L/T2DM vs. PLR-H/T2DM, PLR-L/ Non-T2DM vs. PLR-H/Non-T2DM and PLR-L/ Non-T2DM vs. PLR-H/Non-T2DM vs. PLR-L/T2DM vs. PLR-H/T2DM were presented, and Log-rank test was used to evaluate the statistical significance. When analyzing PLR as a continuous variable, the Log-transformation of PLR was performed (ln [platelet-to-lymphocyte ratio]). Restricted cubic spline (RCS) with 4 knots (5^th^, 35^th^, 65^th^, and 95^th^ percentiles) in the fully adjusted model was used to estimate the dose-response relationship between PLR and the risk of outcomes. Nonlinearity was tested using the likelihood ratio test.

In the muti-COX regression model, PLR-L/Non-T2DM and PLR-L/ T2DM were referenced. Adjusted cofounding factors include age, male sex, body mass index (BMI), hypertension, dyslipidemia, smoking history, previous MI, previous PCI/CABG, previous stroke, previous PAD, Acute coronary syndrome (ACS), LM/three-vessel disease, chronic total occlusion (CTO), moderate to severe calcification, procedural success, number of treated vessels, number of stents, TC, LDL-C, hs-CRP, creatine, left ventricular ejection fraction (LVEF), DAPT, β blocker, and statins. The hazard ratio (HR) and 95% confidence interval (CI) were presented. Sub-group analysis was further performed in 6 different subsets (sex [male or female], age [< 65 or ≥ 65 years], BMI [<25.0 or ≥25.0 kg/m^2^], smoking status [never or former and current], hypertension [yes or no], and ACS [yes or no]), which was presented with a forest plot. Sensitivity analyses excluding patients who presented MACCE within the first 30 days of follow-up, did not receive DAPT and did not receive successful PCI were progressively conducted based on the fully adjusted model.

Statistically, significance was determined when two-sided α was less than 0.05. All the statistical analyses were done using the SPSS version 27.0 (IBM Corporation, Armonk, NY) and RStudio software (version 2021.09.0; http://www.rstudio.org/).

## 3. Results

### 3.1. Baseline characteristics

Generally, 8831 consecutive CAD patients (58.39 ± 10.27 years, 77.0% male) who received PCI with DES implantation were included in this study. The baseline characteristics of four groups based on PLR levels and glycemic metabolism status were listed in **Table 1**. Compared with diabetic patients with a higher level of PLR, those in the other three groups were older and more likely to be female, with a higher prevalence of hypertension, dyslipidemia, previous revascularization, previous stroke, and previous PAD. Laboratory test results including PLR and CK-MB were significantly higher in the PLR-H/T2DM. For coronary procedural characteristics, individuals in the PLR-H/T2DM group were more likely to have a higher SYNTAX score and LM/three-vessel disease when compared with those in the other 3 groups. Besides, the comparisons in baseline characteristics between the PLR-low and PLR-high groups were also performed and presented in **Table S1**.

**Table 1.** Baseline characteristics by different levels of PLR in patients with or without T2DM.

| Variable | PLR-L-NT2D<br>M<br>(N =3015) | PLR-H-NT2D<br>M<br>(N =3149) | PLR-L-T2DM<br>(N =1404) | PLR-H-T2DM<br>(N =1263) | P value |
| --- | --- | --- | --- | --- | --- |
| **Baseline characteristics** |  |  |  |  |  |
| Age, years | 57.79 ± 10.28 | 58.54 ± 10.56 | 58.11 ± 9.70 | 59.71 ± 9.94 | < 0.001 |
| Male sex | 2468 (81.9) | 2230 (74.0) | 1090 (77.6) | 909 (72.0) | < 0.001 |
| BMI, kg/m<sup>2</sup> | 26.19 ± 3.21 | 25.35 ± 3.18 | 26.56 ± 3.15 | 26.02 ± 3.37 | < 0.001 |
| Hypertension | 1834 (60.8) | 2009 (63.8) | 934 (66.5) | 924 (73.2) | < 0.001 |
| Dyslipidemia | 1988 (65.9) | 1942 (61.7) | 1018 (72.5) | 959 (75.9) | < 0.001 |
| Smoking history | 1883 (62.6) | 1667 (53.0) | 815 (58.1) | 652 (51.7) | < 0.001 |
| Previous MI | 615 (20.4) | 521 (16.5) | 292 (20.8) | 232 (18.4) | < 0.001 |
| Previous PCI/CABG | 709 (23.5) | 698 (22.2) | 385 (25.2) | 355 (28.1) | < 0.001 |
| Previous PAD | 209 (6.9) | 205 (6.5) | 124 (8.8) | 138 (10.9) | < 0.001 |
| Previous stroke | 275 (9.1) | 297 (9.4) | 169 (12.0) | 175 (13.9) | < 0.001 |
| LVEF | 63.57 ± 6.87 | 63.51 ± 6.79 | 63.61 ± 6.30 | 62.90 ± 7.83 | 0.048 |
| Clinical presentation |  |  |  |  |  |
| CCS | 1300 (43.1) | 1185 (37.6) | 663 (47.2) | 520 (41.2) | < 0.001 |
| ACS | 1715 (56.9) | 1964 (62.4) | 741 (52.8) | 743 (58.8) | < 0.001 |
| Signs |  |  |  |  |  |
| SBP | 126.70 ± 17.03 | 126.69 ± 16.96 | 128.17 ± 16.89 | 128.69 ± 17.01 | < 0.001 |
| DBP | 77.78 ± 10.63 | 77.94 ± 11.08 | 77.48 ± 10.04 | 77.03 ± 10.33 | 0.311 |
| **Laboratory tests** |  |  |  |  |  |
| Hgb, g/L | 146.48 ± 14.65 | 143.11 ± 15.08 | 144.78 ± 14.36 | 140.53 ± 15.85 | < 0.001 |
| Platelet, 10<sup>9</sup>/L | 186.76 ± 44.92 | 227.29 ± 56.51 | 184.33 ± 41.58 | 225.44 ± 62.32 | < 0.001 |
| White blood cell, 10<sup>9</sup>/L | 6.87 ± 1.58 | 6.26 ± 1.58 | 7.06 ± 1.42 | 6.47 ± 1.61 | < 0.001 |
| Lymphocyte, 10<sup>9</sup>/L | 2.23 ± 0.57 | 1.59 ± 0.44 | 2.30 ± 0.59 | 1.60 ± 0.45 | < 0.001 |
| CK-MB, IU/L | 11.82 ± 8.40 | 11.35 ± 5.74 | 11.55 ± 5.37 | 13.26 ± 21.11 | 0.044 |
| c-TnI, ng/ml | 0.09 ± 0.43 | 0.15 ± 0.79 | 0.06 ± 0.35 | 0.19 ± 1.10 | 0.093 |
| Lp(a), mg/dL | 27.94 ± 28.10 | 31.18 ± 29.94 | 25.01 ± 25.39 | 29.32 ± 29.09 | < 0.001 |
| TC, mmol/L | 4.23 ± 1.08 | 4.30 ± 1.04 | 4.07 ± 1.00 | 4.10 ± 1.08 | < 0.001 |
| HDL-C, mmol/L | 1.04 ± 0.27 | 1.09 ± 0.30 | 1.00 ± 0.28 | 1.02 ± 0.25 | < 0.001 |
| LDL-C, mmol/L | 2.54 ± 0.92 | 2.60 ± 0.89 | 2.37 ± 0.81 | 2.42 ± 0.90 | < 0.001 |
| HbA1c, % | 6.14 ± 0.74 | 6.06 ± 0.54 | 7.84 ± 1.40 | 7.77 ± 1.35 | < 0.001 |
| hs-CRP, mg/L | 2.67 ± 3.28 | 3.02 ± 3.65 | 2.57 ± 3.04 | 3.21 ± 3.92 | < 0.001 |
| Creatinine, umol/L | 76.11 ± 15.30 | 74.14 ± 14.44 | 73.30 ± 14.63 | 73.64 ± 16.02 | < 0.001 |
| **Medications** |  |  |  |  |  |
| DAPT | 2939 (97.5) | 3066 (97.4) | 1378 (98.1) | 1243 (98.4) | 0.098 |
| Aspirin | 2978 (98.8) | 3110 (98.8) | 1396 (99.4) | 1250 (99.0) | 0.195 |
| Clopidogrel | 2974 (98.6) | 3101 (98.5) | 1385 (98.6) | 1253 (99.2) | 0.296 |
| GPIIb/IIIa receptor antagonists | 489 (16.2) | 542 (17.2) | 223 (15.9) | 207 (16.4) | 0.634 |
| β-blocker | 2682 (89.0) | 2811 (89.3) | 1291 (92.0) | 1164 (92.2) | < 0.001 |
| Statins | 2902 (96.3) | 3034 (96.3) | 1343 (95.7) | 1208 (95.6) | 0.543 |
| Nitrate | 2960 (98.2) | 3073 (97.6) | 1372 (97.7) | 1233 (97.6) | 0.411 |
| **Coronary procedural data** |  |  |  |  |  |
| SYNTAX Score | 11.87 ± 7.89 | 11.65 ± 7.69 | 12.57 ± 7.88 | 12.75 ± 8.28 |  |
| LM/three-vessel disease | 1221 (40.5) | 1308 (41.5) | 692 (49.3) | 677 (53.6) | < 0.001 |
| Chronic total occlusion | 490 (16.3) | 481 (15.3) | 210 (15.0) | 194 (15.4) | 0.633 |
| Bifurcation | 528 (17.5) | 512 (16.3) | 239 (17.0) | 190 (15.0) | 0.216 |
| Moderate to severe calcification | 422 (16.3) | 458 (14.5) | 232 (16.5) | 226 (17.9) | 0.004 |
| Number of treated vessels | 1.41 ± 0.66 | 1.36 ± 0.59 | 1.45 ± 0.69 | 1.44 ± 0.67 | < 0.001 |
| Number of stents | 1.91 ± 1.05 | 1.80 ± 0.94 | 2.00 ± 1.06 | 1.95 ± 1.07 | < 0.001 |
| Procedural success | 2973 (99.3) | 3103 (99.4) | 1385 (99.2) | 1251 (99.5) | 0.754 |

Values are mean ± standard deviation or n (%). PLR, platelet-to-lymphocyte ratio; BMI, body mass index; MI, myocardial infarction; PCI, percutaneous coronary intervention; CABG, coronary artery bypass grafting; PAD, peripheral artery disease; CCS, chronic coronary syndrome; ACS, acute coronary syndrome; SBP, systolic blood pressure; DBP, diastolic blood pressure; Lp(a), lipoprotein(a); TC, total cholesterol; HDL-C, high-density lipoprotein cholesterol; LDL-C, low-density lipoprotein cholesterol; HbA1c, glycosylated hemoglobin A1c; hsCRP, high-sensitivity C-reactive protein; LVEF, left ventricular ejection fraction; DAPT, dual antiplatelet therapy; LM, left main; DAPT, dual anti-platelet therapy; SYNTAX, synergy between PCI with taxus and cardiac surgery.

**Table 2.** Prognostic value of PLR for risk of MACCE, individual components and stent thrombosis.

| Groups | Events (%) | Univariable |  | Multivariable* |  |
| --- | --- | --- | --- | --- | --- |
|  |  | HR (95%CI) | *P* value | HR (95%CI) | *P* value |
| MACCEs |  |  |  |  |  |
| PLR-L | 293 (6.6) | Ref | - | Ref | - |
| PLR-H | 370 (8.4) | 1.29 (1.11-1.50) | 0.001 | 1.26 (1.07-1.47) | 0.005 |
| All-cause mortality |  |  |  |  |  |
| PLR-L | 32 (0.7) | Ref | - | Ref | - |
| PLR-H | 69 (1.6) | 2.20 (1.45-3.35) | < 0.001 | 1.87 (1.21-2.89) | 0.005 |
| MI |  |  |  |  |  |
| PLR-L | 40 (0.9) | Ref | - | Ref | - |
| PLR-H | 57 (1.3) | 1.46 (0.97-2.18) | 0.069 | 1.44 (0.95-2.18) | 0.090 |
| TVR |  |  |  |  |  |
| PLR-L | 192 (4.3) | Ref | - | Ref | - |
| PLR-H | 222 (5.0) | 1.18 (0.97-1.43) | 0.099 | 1.18 (0.97-1.45) | 0.098 |
| Stroke |  |  |  |  |  |
| PLR-L | 58 (1.3) | Ref | - | Ref | - |
| PLR-H | 73 (1.7) | 1.30 (0.92-1.83) | 0.139 | 1.24 (0.87-1.77) | 0.241 |
| Stent thrombosis |  |  |  |  |  |
| PLR-L | 23 (0.5) | Ref | - | Ref | - |
| PLR-H | 52 (1.2) | 2.31 (1.41-3.77) | < 0.001 | 2.29 (1.39-3.79) | 0.001 |

\*Adjusted for age, male sex, BMI, hypertension, dyslipidemia, smoking history, previous MI, previous PCI/CABG, previous stroke, previous PAD, ACS, SYNTAX score, LM/three-vessel disease, chronic total occlusion, moderate to severe calcification, procedural success, number of treated vessels, number of stents, TC, LDL-C, hsCRP, creatine, LVEF, DAPT, $\beta$ blocker and statins.
PLR, platelet-to-lymphocyte ratio; MACCEs, major adverse cardiac cerebral events; MI, myocardial infarction; TVR, target vessel revascularization; HR, hazard ratio; CI, confidence interval; Ref, reference.

### 3.2. PLR levels and the risk of MACCE in the whole cohort

During a median follow-up of 2.4 years (interquartile range: 2.2-2.6 years), 101 (1.1%) all-cause deaths, 97 (1.1%) MI, 131 (1.5%) strokes, 414 (4.7%) TVR, and 663 (7.5%) MACCEs were recorded. **Table 3** displayed the comparison of clinical outcomes risks between the PLR-H and PLR-L groups. In multivariable analysis after adjusting for confounding factors, the PLR-H group was at significantly higher risk of MACCE (HR: 1.26, 95%CI: 1.07 to 1.47, *P* = 0.005), and all-cause death (HR: 1.87, 95%CI: 1.21 to 2.89, *P* = 0.005). Consistently, the K-M analysis curves also revealed a higher risk of MACCE in patients with PLR-H compared with those with PLR-L (log-rank *P* = 0.001, **Figure 1A**).

**Figure 1.**
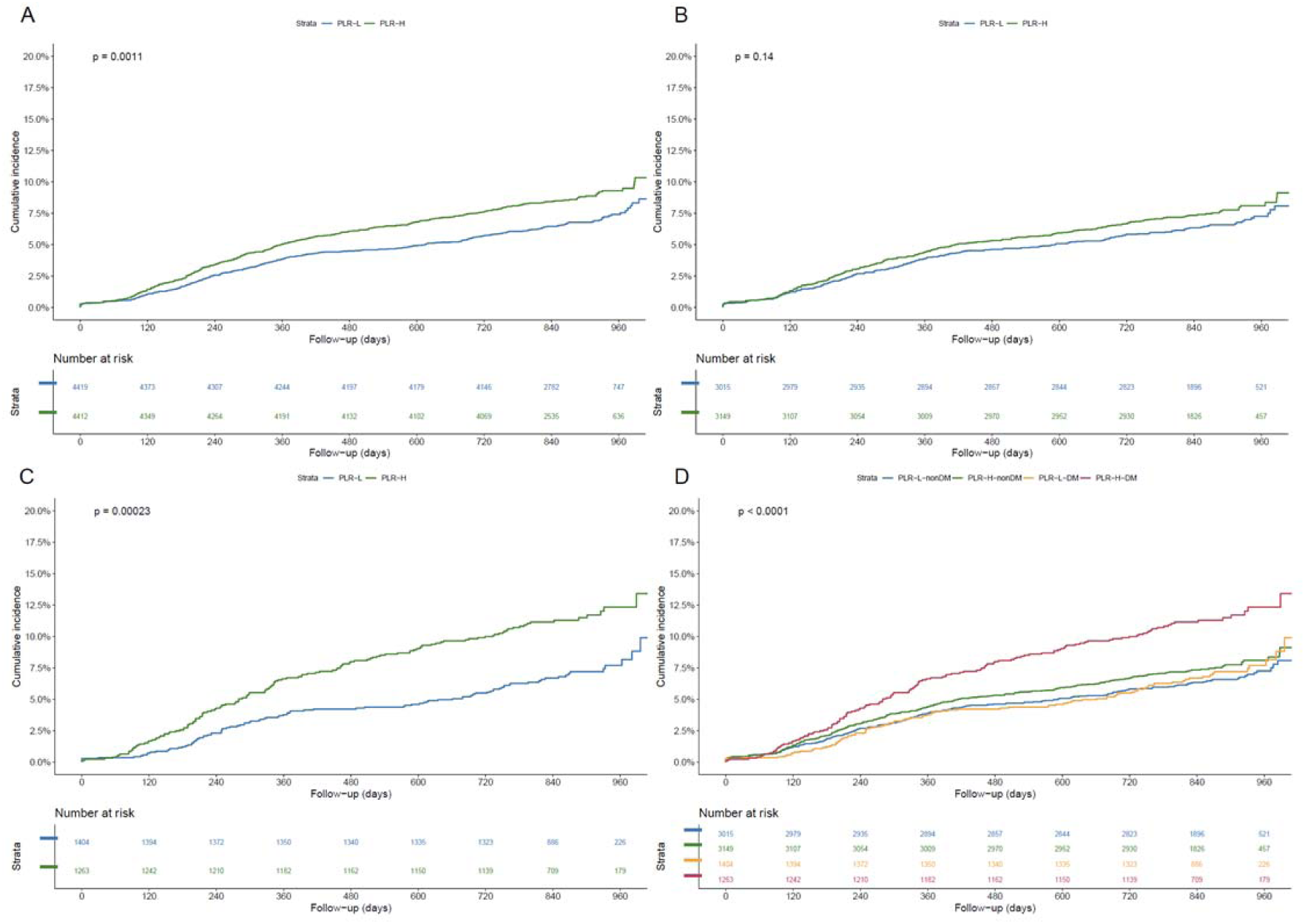
**Kaplan–Meier curves for cumulative incidence of MACCE**, according to different PLR levels (A), different PLR levels in non-T2DM patients (B) and T2DM patients (C), and four risk groups (PLR-L/non-T2DM, PLR-H/non-T2DM, PLR-L/T2DM and PLR-H/T2DM) (D). PLR-L, low level of platelet to lymphocyte ratio (< 107.0); PLR-H, high level of platelet to lymphocyte ratio (≥ 107.0); T2DM, type 2 diabetes mellitus.

**Table 3.**
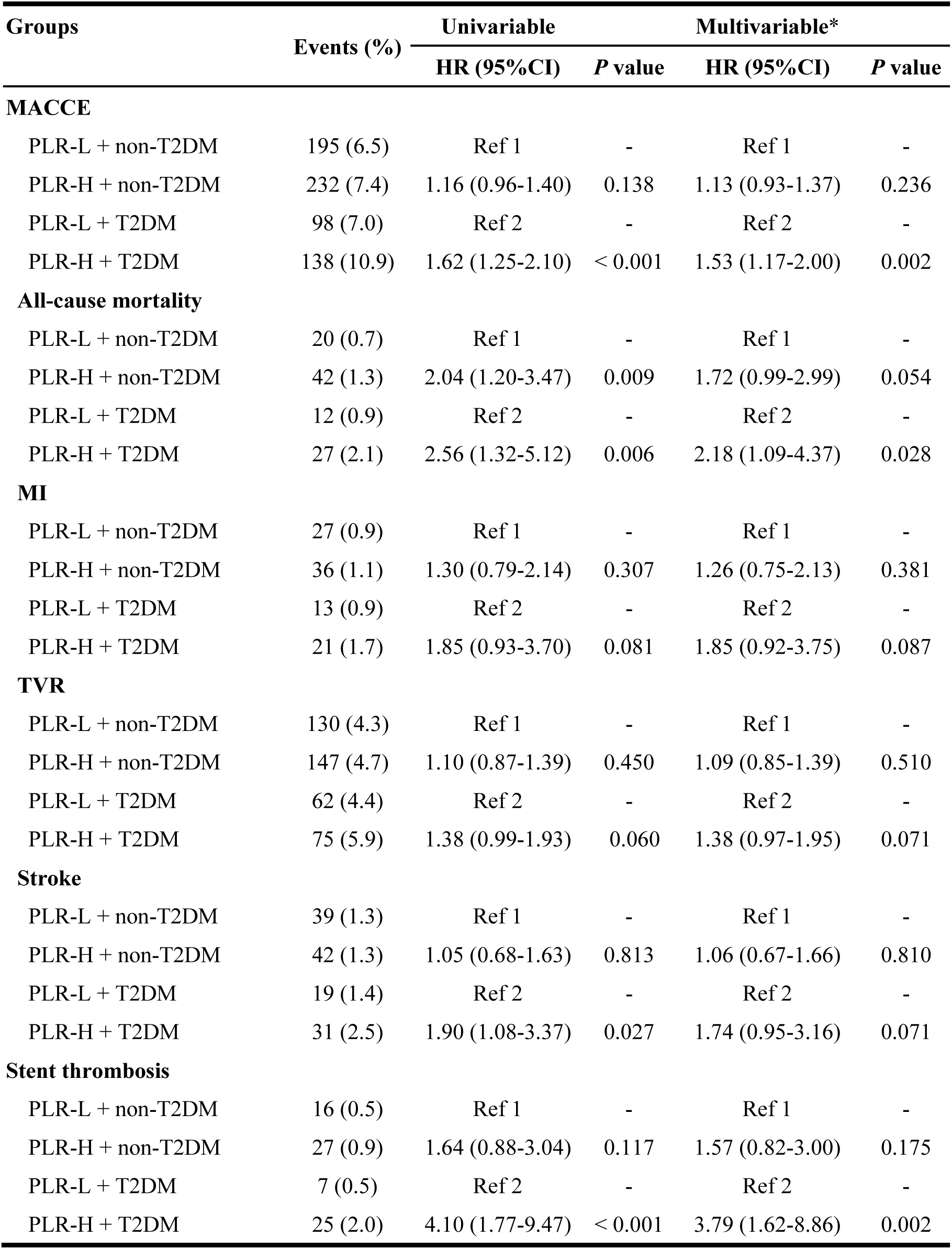
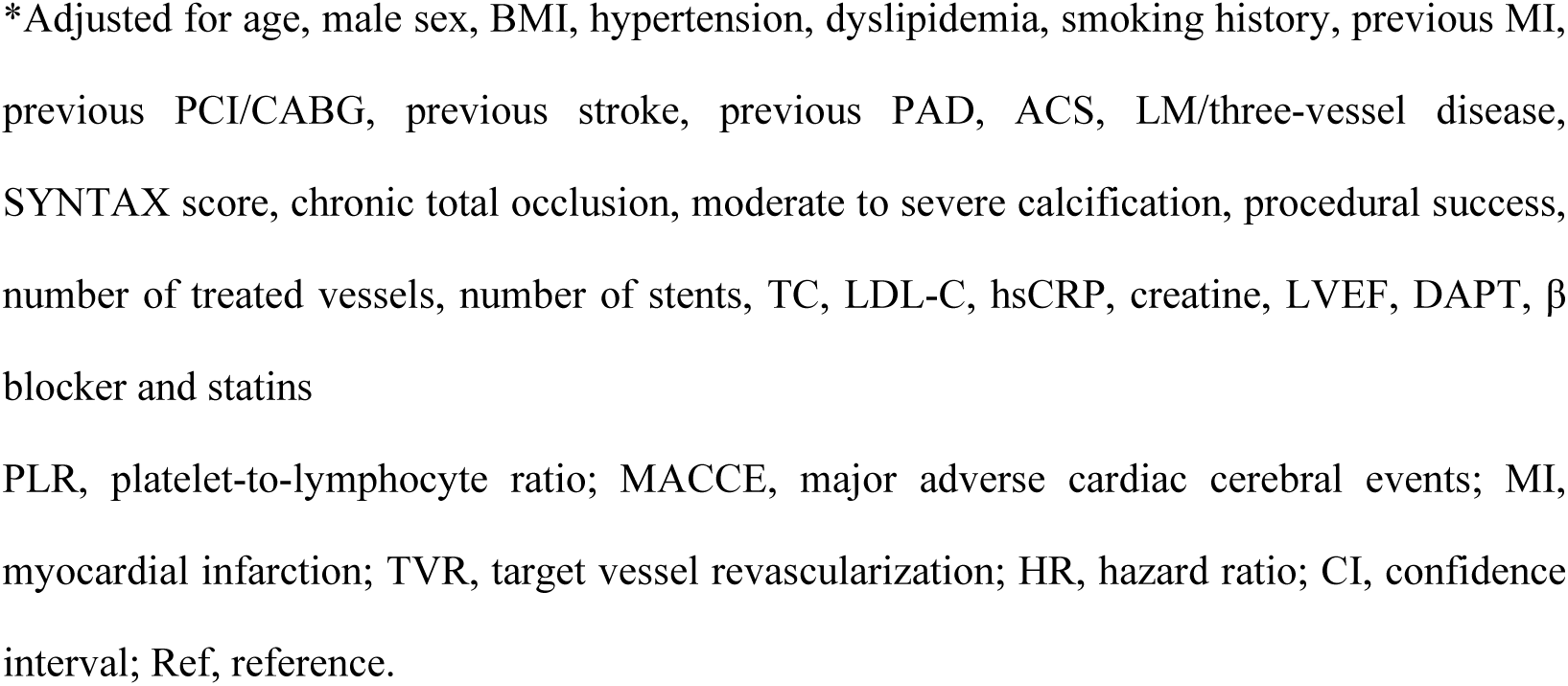
Prognostic value of combined status of T2DM and PLR for risk of MACCEs, individual components and stent thrombosis.

| Groups | Events (%) | Univariable |  | Multivariable* |  |
| --- | --- | --- | --- | --- | --- |
|  |  | HR (95%CI) | *P* value | HR (95%CI) | *P* value |
| MACCE |  |  |  |  |  |
| PLR-L + non-T2DM | 195 (6.5) | Ref 1 | - | Ref 1 | - |
| PLR-H + non-T2DM | 232 (7.4) | 1.16 (0.96-1.40) | 0.138 | 1.13 (0.93-1.37) | 0.236 |
| PLR-L + T2DM | 98 (7.0) | Ref 2 | - | Ref 2 | - |
| PLR-H + T2DM | 138 (10.9) | 1.62 (1.25-2.10) | < 0.001 | 1.53 (1.17-2.00) | 0.002 |
| All-cause mortality |  |  |  |  |  |
| PLR-L + non-T2DM | 20 (0.7) | Ref 1 | - | Ref 1 | - |
| PLR-H + non-T2DM | 42 (1.3) | 2.04 (1.20-3.47) | 0.009 | 1.72 (0.99-2.99) | 0.054 |
| PLR-L + T2DM | 12 (0.9) | Ref 2 | - | Ref 2 | - |
| PLR-H + T2DM | 27 (2.1) | 2.56 (1.32-5.12) | 0.006 | 2.18 (1.09-4.37) | 0.028 |
| MI |  |  |  |  |  |
| PLR-L + non-T2DM | 27 (0.9) | Ref 1 | - | Ref 1 | - |
| PLR-H + non-T2DM | 36 (1.1) | 1.30 (0.79-2.14) | 0.307 | 1.26 (0.75-2.13) | 0.381 |
| PLR-L + T2DM | 13 (0.9) | Ref 2 | - | Ref 2 | - |
| PLR-H + T2DM | 21 (1.7) | 1.85 (0.93-3.70) | 0.081 | 1.85 (0.92-3.75) | 0.087 |
| TVR |  |  |  |  |  |
| PLR-L + non-T2DM | 130 (4.3) | Ref 1 | - | Ref 1 | - |
| PLR-H + non-T2DM | 147 (4.7) | 1.10 (0.87-1.39) | 0.450 | 1.09 (0.85-1.39) | 0.510 |
| PLR-L + T2DM | 62 (4.4) | Ref 2 | - | Ref 2 | - |
| PLR-H + T2DM | 75 (5.9) | 1.38 (0.99-1.93) | 0.060 | 1.38 (0.97-1.95) | 0.071 |
| Stroke |  |  |  |  |  |
| PLR-L + non-T2DM | 39 (1.3) | Ref 1 | - | Ref 1 | - |
| PLR-H + non-T2DM | 42 (1.3) | 1.05 (0.68-1.63) | 0.813 | 1.06 (0.67-1.66) | 0.810 |
| PLR-L + T2DM | 19 (1.4) | Ref 2 | - | Ref 2 | - |
| PLR-H + T2DM | 31 (2.5) | 1.90 (1.08-3.37) | 0.027 | 1.74 (0.95-3.16) | 0.071 |
| Stent thrombosis |  |  |  |  |  |
| PLR-L + non-T2DM | 16 (0.5) | Ref 1 | - | Ref 1 | - |
| PLR-H + non-T2DM | 27 (0.9) | 1.64 (0.88-3.04) | 0.117 | 1.57 (0.82-3.00) | 0.175 |
| PLR-L + T2DM | 7 (0.5) | Ref 2 | - | Ref 2 | - |
| PLR-H + T2DM | 25 (2.0) | 4.10 (1.77-9.47) | < 0.001 | 3.79 (1.62-8.86) | 0.002 |

\*Adjusted for age, male sex, BMI, hypertension, dyslipidemia, smoking history, previous MI, previous PCI/CABG, previous stroke, previous PAD, ACS, LM/three-vessel disease, SYNTAX score, chronic total occlusion, moderate to severe calcification, procedural success, number of treated vessels, number of stents, TC, LDL-C, hsCRP, creatine, LVEF, DAPT, $\beta$ blocker and statins PLR, platelet-to-lymphocyte ratio; MACCE, major adverse cardiac cerebral events; MI, myocardial infarction; TVR, target vessel revascularization; HR, hazard ratio; CI, confidence interval; Ref, reference.

When analyzing PLR as a continuous variable, results showed that a per 1 unit increase in PLR-ln was associated with a 49% increase in the risk of MACCE (95%CI: 1.18 to1.87, *P* = 0.001) **(Table S2)**. RCS analysis further determined a linear relationship between PLR and the risk of MACCE in the whole cohort (P for non-linearity = 0.071) **(Figure S2)**.

### 3.3. PLR levels and the risk of MACCE in patients with and without T2DM

The incidence of MACCE in PLR-H with T2DM or non-T2DM and PLR-L with T2DM or non-T2DM group was 14.3% (178/1263), 10.6% (337/3149), 11.1% (158/1404), 10.6% (318/3015), respectively. Risks of MACCE were compared in four different risk groups by the Cox regression model, using the PLR-L-non-T2DM and PLR-L-DM groups as reference 1 and 2, respectively **(Table 3)**. Compared with reference 1, patients in the PLR-H/non-T2DM were at significantly higher risk of all-cause mortality in univariable analysis (HR: 2.04, 95%CI: 1.20 to 3.47, *P* = 0.009), while no significant difference was presented in multivariable analysis. When it came to referencing 2, patients with PLR-H/T2DM showed a significantly higher risk of MACCE (HR: 1.62, 95%CI: 1.25 to 2.10, *P* < 0.001), all-cause mortality (HR: 2.56, 95%CI: 1.32 to 5.12, *P* = 0.006), stroke (HR: 1.90, 95%CI: 1.08 to 3.37, *P* = 0.027) and stent thrombosis (HR: 4.10, 95%CI: 1.77 to 9.47, *P* < 0.001). In multivariable analysis after adjusting for confounding factors, increased risk of MACCE (HR: 1.53, 95%CI: 1.17 to 2.00, *P* = 0.002), all-cause mortality (HR: 2.18, 95%CI: 1.09 to 4.37, *P* = 0.028) and stent thrombosis (HR: 3.79, 95%CI, 1.62 to 8.86, *P* = 0.002) were still significant in PLR-H/T2DM group. When it came to the continuous analysis, with PLR-Ln increasing per 1 unit, the risk of MACCE and stent thrombosis increased by 99% (95%CI: 1.37 to 2.89, *P* < 0.001) in the T2DM cohort with great significance (**Table S2)**. However, this significant association was not found in the non-T2DM cohort.

### 3.4. PLR levels and the risk of stent thrombosis

During the follow-up, there were 75 (0.85%) cases of stent thrombosis were recorded. In the whole cohort study, K-M plots showed that patients in the PLR-H group presented a significantly higher risk of stent thrombosis compared with patients in PLR-L (**Figure 2A**). This phenomenon was still significant in a patient with T2DM, but not remarkable in patients with non-T2DM **(Figure 2B, C)**, and patients with both PLR-H and T2DM held the highest risk (**Figure 2D**). In Cox regression analyses, the risk of stent thrombosis was significantly higher in the PLR-High group when compared with the PLR-Low group in the T2DM cohort **(Table 3)**. When it came to the continuous analysis, the risk of stent thrombosis increased significantly with the increase in PLR-Ln levels in the T2DM cohort **(Table S2)**.

**Figure 2.**
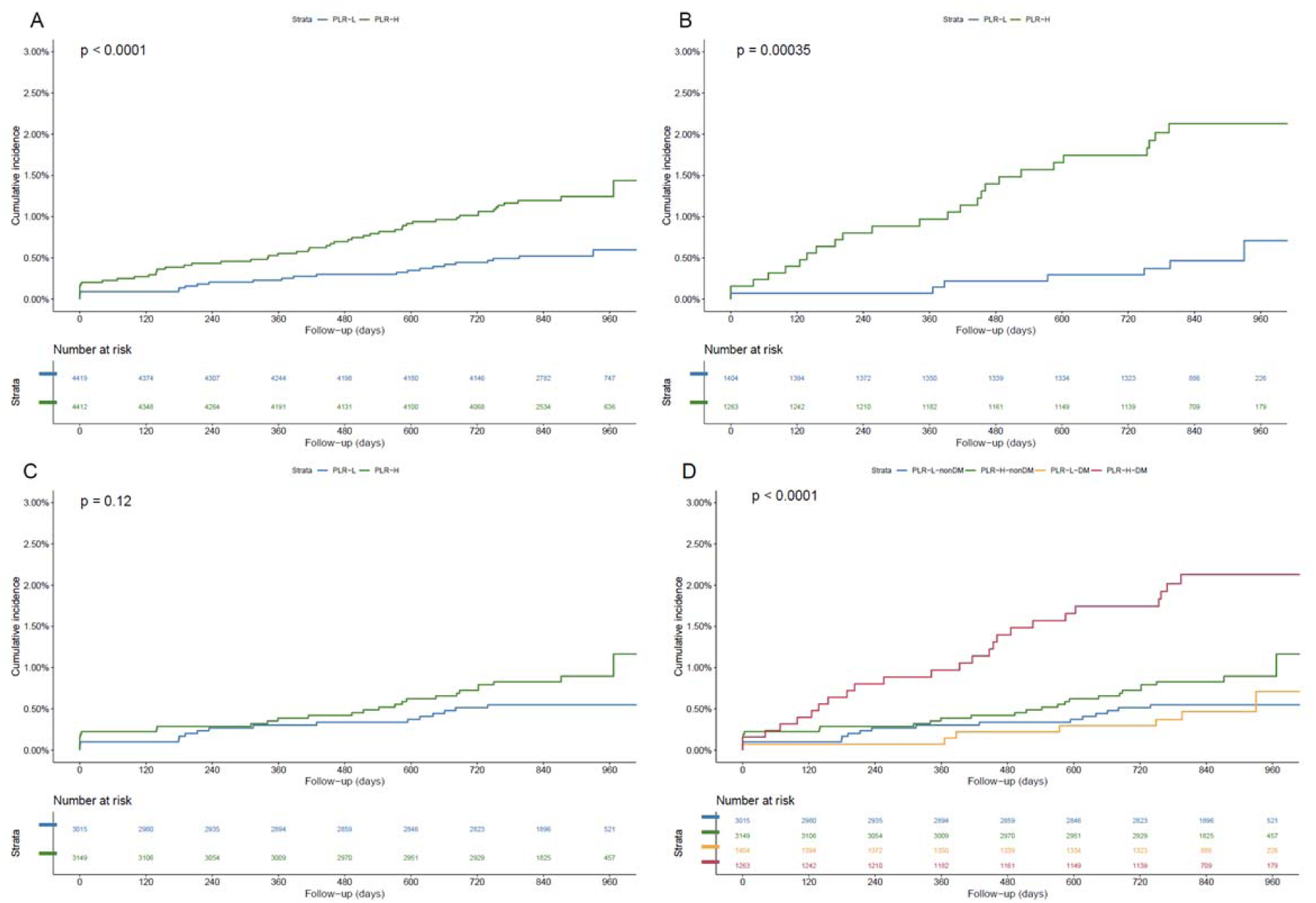
**Kaplan–Meier curves for cumulative incidence of stent thrombosis, according to different PLR levels (A), and four risk groups (PLR-L/non-T2DM, PLR-H/non-T2DM, PLR-L/T2DM and PLR-H/T2DM) (D).** PLR-L, low level of platelet to lymphocyte ratio (< 107.0); PLR-H, high level of platelet to lymphocyte ratio (≥ 107.0); T2DM, type 2 diabetes mellitus.

We further performed the adjusted Cox regression model for the subtypes of stent thrombosis in the whole cohort and DM cohort **(Table S3)**. Results showed that VLST occupied the largest proportion (52%) and was the only subtype whose risk was significantly positively associated with the PLR-Ln levels in T2DM (HR 2.04, 95%CI: 1.34 to 5.64, *P* = 0.008).

### 3.5. Subgroup and sensitivity analysis

Subgroup analysis was performed to evaluate the association of risk groups and MACCE and stent thrombosis across different subgroups, including age, sex, BMI, hypertension, smoking status, and ACS **(Figure S3, Figure S4, and Table S4, S5)**. There was a significant interaction between risk groups and MACCE and stent thrombosis by age of 65 years (P for interaction = 0.002 and 0.031, respectively), whereas the consistent association of risk groups with MACCE risk was observed in patients with different sex, BMI, hypertension-status, smoking-status and clinical presentation (P for interaction > 0.05) after multiple adjustments. Sensitivity analysis for excluding patients who presented MACCE within the first 30 days of follow-up, received unsuccessful PCI, and did not receive DAPT was further performed with the outcomes of MACCE and stent thrombosis, and no change in conclusions was found in three cohorts **(Table S6)**. Sensitivity analysis for individuals of MACCE was presented in **Table S7-S9**.

## 4. Discussion

In this real-world, large-scale, prospective, observational cohort study, the prognosis of diabetic and non-diabetic CAD patients undergoing PCI with different levels of PLR was investigated. The major findings of this study are as follows: (1) high PLR level is significantly associated with the increased risk of MACCE in patients undergoing PCI with T2DM; (3) high PLR level is an independent predictor for stent thrombosis in patients with T2DM but not in those without DM, and the VLST is the main subtype; (4) the correlation between PLR-ln and the risk of MACCE is a positive linear type, and the risk of MACCE and stent thrombosis elevated significantly with per 1 unit increasing in PLR-ln in patients with CAD and T2DM; (5) patients combined with both T2DM and high PLR levels presented the worst prognosis and the highest risk of stent thrombosis. These results were not changed after performing subgroup and sensitivity analysis.

PLR is an easy-to-obtained inflammatory biomarker calculated as the ratio of the platelet to lymphocyte count. Previous studies had applied PLR in several inflammatory disorder conditions, including solid tumors ^17^, arthritis ^18^, and acute kidney injury ^19^. These studies indicated PLR could be a useful biomarker to indicate the disease activity, treatment reaction, and prognosis. For cardiovascular disease, several studies have investigated the role of PLR in patients with ACS or CCS. Cetin et al.^20^ recruited 1938 acute STEMI patients with primary PCI to investigate the prognostic value of PLR evaluated before PCI with the primary endpoint of MACE (stent thrombosis, nonfatal myocardial infarction, and cardiovascular mortality). Results showed that higher PLR was associated with increased risk of in-hospital MACCE (>143 vs. ≤143; HR: 2.434; 95%CI, 1.528 to 3.878) and long-term MACCE (>147 vs. ≤147; HR: 2.810, 95%CI: 2.014 to 3.920). Besides, Azab et al. investigated the predictive value of PLR for 4-year mortality in 619 NSTEMI patients and found elevated admission PLR has associated with a higher risk of 4-year mortality (>176 vs. ≤176; HR: 1.62; 95%CI, 1.15 to 2.26) ^21^. Meanwhile, Açar et al. ^9^ investigated the relationship between the PLR and coronary collateral circulation (CCC) in CCS patients. They found per an elevated 10-unit PLR was associated with poor CCC (odds ratio, 1.48; 95%CI, 1.33 to 1.65). These studies indicated PLR had well predictive ability in patients with CAD. Consistently, our research revealed that a high PLR level (>107.0) held great predictive value for the poor prognosis in patients undergoing PCI. Further, unlike previous studies, we demonstrated a linear relationship between PLR level and MACCE risk for the first time and revealed that a per 1 unit increase in PLR-ln could bring a significant elevation in MACCE risk.

PLR-related prothrombotic status and high systemic inflammation were indicated as two main pathways conferring cardiotoxicity, as we found that the risk of stent thrombosis was significantly higher in PLR-H patients than those with PLR-L, and high levels of inflammatory biomarkers were presented in patients with high PLR. For the underlying mechanisms, a high level of platelets was associated with relative thrombocytosis and megakaryocytic proliferation which resulted in atherosclerosis and thrombotic events ^22, 23^. Besides, high PLR was also associated with increased release of pro-inflammatory cytokines, including interleukin-1, interleukin-6, and tumor necrotizing factor-α, which exacerbated inflammatory burden, and elevated myocardium damage and remodeling ^24, 25^. Last, decreased lymphocyte, which has been indicated as a predictor for the poor prognosis in ST-segment elevated MI patients ^26^, was shown to confer increasing cardiomyocyte apoptosis under the high inflammatory burden, which further exaggerated myocardial injury ^27^.

The predictive value of high PLR with poor prognosis in CAD patients with T2DM was also determined for the first time. To our knowledge, high PLR was demonstrated as a predictor for poor prognosis in patients with T2DM, as previous research showed that high PLR was associated with the increased risk of several complications like diabetic nephropathy ^12^, diabetic peripheral neuropathy ^28^, diabetic neurogenic bladder ^29^, cognitive decline ^13^ and carotid plaque ^30^. Consistent with these studies, CAD patients in the T2DM-PLR-H group showed the highest risk of all clinical outcomes and the worst prognosis. For the mechanisms, it hypothesizes that the status of hyperglycemia further exacerbates the pro-inflammatory and pro-thrombotic roles of PLR we discussed above, as it has been widely revealed that elevated blood glycose upregulated the inflammatory burden and enhanced the activity of platelet ^31, 32^. Interestingly, high-PLR related poor prognosis was not significant in CAD patients with non-DM. This phenomenon might be attributed to the lack of potential cardiotoxic interactions between hyperglycemia and PLR, such as increased platelet activity induced by high glucose. However, there is no related investigations so far, and further studies are expected to focus on non-DM people and present potential explanations.

Another breakthrough in our study was the first to reveal that PLR was an independent risk factor for 2-year stent thrombosis after PCI in T2DM patients. Stent thrombosis was still a tough issue for patients who underwent PCI and was associated with mortality between 5% and 45% ^33^. Known risk factors for stent thrombosis included premature discontinuation, genetic polymorphisms (for AST or SAST), peripheral arterial occlusive disease (for LST or VLST), DM, malignant disease, smoking (for all types) ^33, 34^. The association of high PLR levels with the risk of coronary stent thrombosis was first explored in our study, and the results presented a positive correlation. As for the underlying mechanism, high platelet levels-induced prothrombotic status, which is further exacerbated in hyperglycemia, is hypothesized as the main contributor. To be noticed, the majority of these stent-thrombosis cases showed as the type of VLST. For this phenomenon, we hypothesized that common treatment (97.7%) of 1-year DAPT in these patients was a potential reason, as a standard DAPT could attenuate the thrombotic status brought by high levels of platelet, which was largely reduced by the common shift from DAPT to aspirin alone after 1 year of implantation. Similar to MACCE, high PLR levels-related high risk of stent thrombosis was also only significant in patients with T2DM, but not remarkable in patients without DM. Since DM is a conventional risk factor for stent thrombosis, and increased platelet activity was demonstrated in high glucose conditions, it was believed that there was a co-joint effect of PLR and hyperglycemia contributing to stent thrombosis.

This study was the first prospective cohort study to investigate the interaction between high PLR levels with the risk of poor prognosis and stent thrombosis for CAD patients under different glycemic statuses. However, there still existed some limitations. First, even though the present study adjusted for a considerable number of potential confounding factors, it was hard to fully adjusted all confounding factors due to the limitation of data collection and the nature of the observational study design. Second, data regarding dynamic changes in PLR and the glycemic status during follow-up are unavailable. Thirdly, the ethnic composition of this study was single, in which only Chinese patients were included. It was still unclear whether the conclusion was suitable for other races. Future well-designed prospective studies and randomized control trials are warranted to confirm our findings.

## 5. Conclusions

PLR is a novel inflammatory biomarker with great predictive values in the poor prognosis and incidence of stent thrombosis in CAD patients with T2DM. Therefore, patients undergoing PCI with high PLR levels should be closely monitored in clinics, especially those combined with T2DM

## Data Availability

The datasets used during the current study are available from the corresponding author upon reasonable request.

## List of abbreviations

ACS: acute coronary syndrome
AST: acute stent thrombosis
BMI: body mass index
CAD: coronary artery disease
CCS: Chronic coronary syndrome
CTO: chronic total occlusion
CI: confidence interval
CABG: coronary artery bypass grafting
CCC: coronary collateral circulation
DES: drug-eluting stents
DAPT: dual anti-platelet therapy
hsCRP: high-sensitivity C-reactive protein
HbA1c: glycosylated hemoglobin A1c
K-M: Kaplan-Meier
LDL-C: low-density lipoprotein cholesterol
LM: left main artery disease
LVEF: left ventricular ejection faction
LST: late stent thrombosis
MI: myocardial infarction
MACCE: major adverse cardiovascular and cerebrovascular events
PCI: percutaneous coronary intervention
PLR: platelet-to-lymphocyte ratio
PAD: peripheral artery disease
STEMI: ST-elevation myocardial infarction
SD: standard deviation
SAST: subacute stent thrombosis
T2DM: type 2 diabetes mellitus
TC: total cholesterol
TG: triglyceride
TIMI: thrombolysis in myocardial infarction
VLST: very late stent thrombosis.

## 6 Acknowledgments

The authors would like to thank all the participants of this study.

## 9. Author contribution

KFD, QTD, and YJS conceived the study. YJS, ZYL, and JNH conducted the analyses. YJS, ZYL, JNH were responsible for the interpretation of the data. YJS, ZYL, JNH, KYC, CXS, RZ, and BQS wrote the first draft of the manuscript. YG and KFD revised it. All authors made substantial contributions to iterations and approved the final version. KFD is responsible for the integrity of this work as a whole.

## 11. Supplemental Material

Tables S1-S9

Figure S1-S4

